# Optimizing Communication Strategies for COPD Management: Effectiveness of Educational Video and Pamphlet Interventions

**DOI:** 10.1101/2025.08.28.25334616

**Authors:** Jeenat Mehareen, Sharon Zhu, Jim Johnson, Mohsen Sadatsafavi, Erica Frank

## Abstract

**Objectives:** Risk prediction models are increasingly used at point of care to support personalized treatment decisions. This study created and evaluated two Information, Education, and Communication (IEC) resources to improve public understanding of a risk prediction tool for Chronic Obstructive Pulmonary Disease (COPD) management.

**Methods:** We created a 5-minute video and a pamphlet explaining the burden of COPD and how a prediction model generates quantitative estimates of, and benefit of certain treatments for, exacerbations of the disease. These tools were tested among students and researchers in public health. A patient partner was engaged throughout to ensure the materials were accessible and patient-centered.

**Results:** Twenty-five individuals participated (80% female; 60% aged 25–64). After reviewing the materials, 92% of participants agreed to the statement “I am familiar with the idea of precision medicine approach”. Most (72%) felt they received sufficient information about the tool, and 92% believed such materials could support patient decision. Participants stated that the materials were clear, detailed, and written in plain language. Participants preferred the pamphlet (68%) over the video (44%). Suggestions for improvement included expanding content on how the tool works.

**Conclusions:** The findings of this study provided a better understanding of how to present complex medical information around precision medicine that is accessible and meaningful to diverse audiences. We will improve our materials based on these comments, and continue to make them available at https://resp.core.ubc.ca/show/patient_committee_2025

## Introduction

Risk prediction models are increasingly implemented at point of care as major enablers of personalized treatment decisions.^1,2^ These models can improve medication adherence and health outcomes as patient characteristics are objectively taken into consideration for guiding healthcare decisions.^3,4^ Effective communication between patients and physicians is important when communicating risks calculated from this model and discussing individualized treatment decisions. However, their complexity often makes it difficult for physicians to explain risk estimates during brief clinical encounters. This communication gap can limit the utility of risk prediction models for shared decision-making.

In the management of Chronic Obstructive Pulmonary Disease (COPD), patients are often faced with complex treatment decisions regarding the prevention of the occurrence of exacerbations (also known as “lung attacks”).^5^ Tools that predict exacerbation risk can provide a backbone for better understanding the risk and for shared decision-making. One such tool is the Acute COPD Exacerbation Prediction Tool (ACCEPT)^6^—a risk prediction model that estimates an individual’s risk of exacerbations in the next 12 months given selected individual and clinical features (predictors) and provides treatment recommendations aligned with existing pharmacotherapy guidelines. Under the IMplementing Predictive Analytics towards efficient COPD Treatments (IMPACT) study, ACCEPT is currently being implemented and examined in two tertiary medical hospitals.^7^ Physicians use the tool’s prediction to provide patients with tailored treatment suggestions. These recommendations are typically delivered as an information sheet, which can be further customized to include individualized advice.

Although patient education has been shown to improve quality of life and reduce hospitalizations^8^, the current information sheets focus mainly on presenting patients’ risk status and corresponding medication recommendations. They offer little insight into how predictions are generated or why the tool should be trusted. This lack of information can undermine patient confidence in the model’s outputs and limit engagement with the recommended treatment plan.

To address the communication gap around risk prediction tools, we conducted a study to develop and test two Information, Education, and Communication (IEC) resources. These materials explain the concept of precision medicine and its application in managing COPD exacerbations in lay language, using ACCEPT as an example. As the ongoing IMPACT study evaluates the clinical and cost-effectiveness of ACCEPT, positive results may lead to broader implementation of the tool. We therefore explored next steps by evaluating how best to communicate model-based risk predictions to patients and the public using these IEC resources. The primary goal was to evaluate how these materials increase understanding of risk prediction tools in COPD care.

## Methods

The study received institutional ethics approval from University of British Columbia (H23-00445). The design of the IEC materials was guided by existing evidence on effective health communication.^9^ Research indicates that combining simple language with visuals improves patient engagement and recall, especially for complex topics like pharmacogenomics and precision medicine.^10,11^ Short videos have also been found to enhance comprehension of medical concepts.^12^ Drawing on this evidence, two materials, a video and a pamphlet—were developed.

A patient partner with lived experience of COPD was engaged throughout the project, beginning with feedback on the initial study design and continuing through the co-creation of materials and review of survey questions used for evaluation. His contributions helped refine both content and presentation, ensured that the resources were relevant, accessible, and meaningful for patients managing COPD as well as understandable to a general audience.

The video (*Please contact corresponding author to request access*), approximately five minutes in length, explained the purpose of the ACCEPT tool, how it predicts the risk of future exacerbations, and how this information supports shared decision-making. To ensure accessibility, the video used plain language and deliberate pacing. The script combined two perspectives: a physician (co-author EF) who explained what COPD is, the concept of precision medicine, and how ACCEPT works, and the patient partner (co-author JJ), who shared his experience living with COPD and discussed how tools like ACCEPT could benefit patients.

A complementary pamphlet (*Supplementary Figure A1*) presented the same key information in a brief, easy-to-read format. The content was adapted from publicly available resources, written at an approximately Grade 8 reading level, and organized with clear sections and visuals to improve readability. The patient partner reviewed and provided feedback on the content, layout drafts, and design features to highlight the most relevant information and improve readability.

Finally, graduate students and researchers in Public Health at the University of British Columbia reviewed both IEC resources and completed the survey, which captured demographics and assessed comprehension, satisfaction, and format preference.

Descriptive statistics were used to summarize participant demographics and feedback on IEC materials. For paired comparisons of ordinal responses (e.g., Likert-scale ratings) between the video and pamphlet formats, the Wilcoxon signed-rank test was used to assess differences in participant perceptions (e.g., clarity, visual engagement, and overall preference). A p-value of <0.05 was considered statistically significant. All analyses were conducted using Microsoft Excel and Rstudio.

## Results

Twenty-five individuals participated in the study (80% female, 60% aged 25-64). 56% were public health students (*Table 1*). *Table 2* provides the participants’ level of agreement regarding the IEC materials. After evaluating both the video and pamphlet, 92% of participants agreed or strongly agreed that they became familiar with the benefits of precision medicine towards COPD management. 72% reported receiving sufficient information about the prediction tool (ACCEPT), while 92% believed similar materials could facilitate conversations about precision medicine applications for disease management among patients. Regarding the clarity and accessibility of the materials, 92% agreed or strongly agreed that the content was easy to understand, and 96% felt that the language used was appropriate and accessible.

**Table 1:**
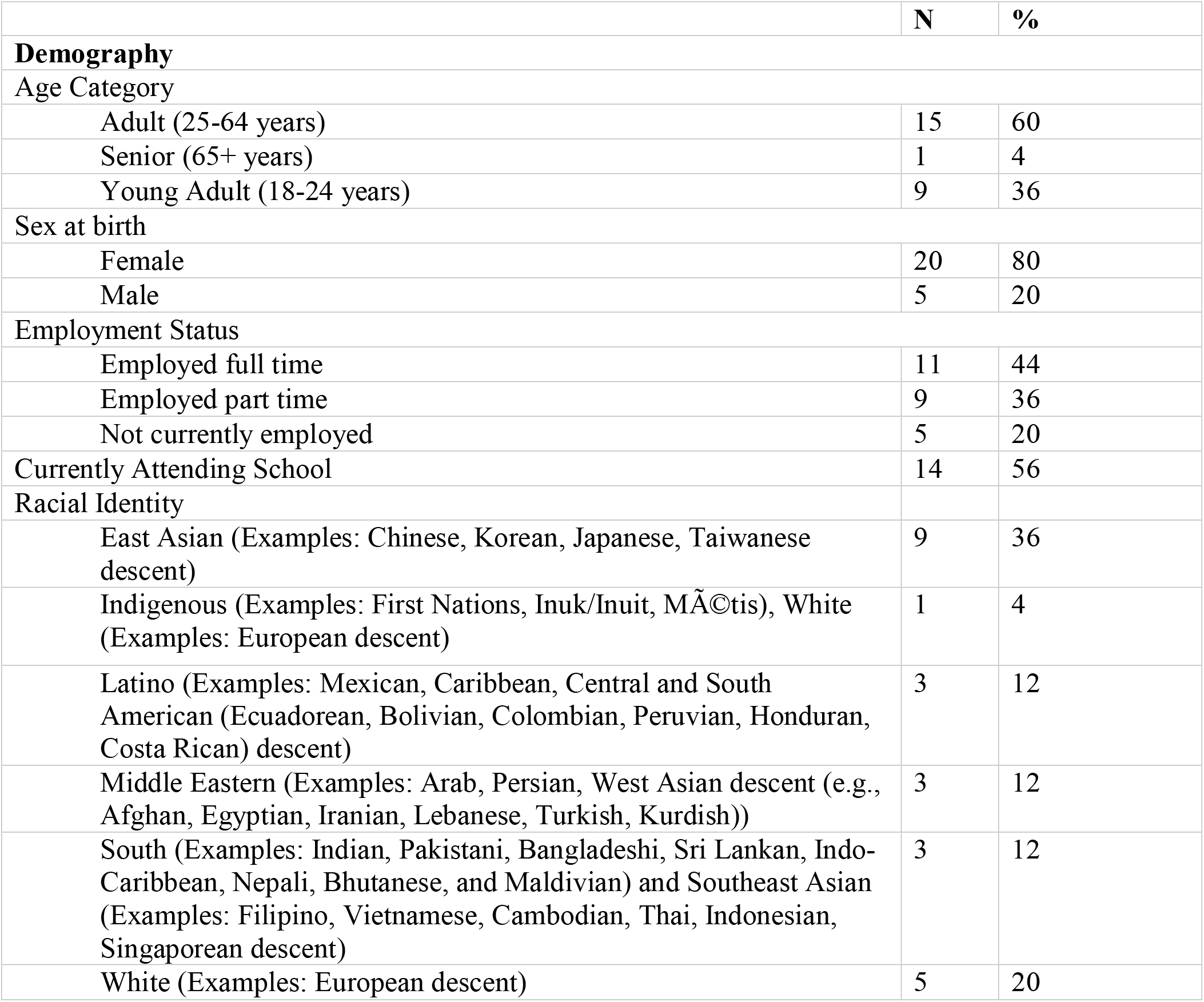
Participant demographics.

**Table 2:** Participants’ feedback on IEC materials.

|  | N | % |
| --- | --- | --- |
| “I am familiar concept of precision medicine” |  |  |
| Disagree | 2 | 8 |
| Neither agree nor disagree | 5 | 20 |
| Agree | 12 | 48 |
| Strongly agree | 6 | 24 |
| “After going through the materials, I am familiar with the idea of precision medicine approach in the management of COPD” |  |  |
| Neither agree nor disagree | 2 | 8 |
| Agree | 14 | 56 |
| Strongly agree | 9 | 36 |
| “These materials provided me with enough information about how a precision medicine tool (ACCEPT) works” |  |  |
| Disagree | 2 | 8 |
| Neither agree nor disagree | 5 | 20 |
| Agree | 13 | 52 |
| Strongly agree | 5 | 20 |
| “The material was engaging and presented the message in a straightforward way.” |  |  |
| Agree | 18 | 72 |
| Strongly Agree | 7 | 28 |
| “The materials were sufficiently explained using plain language.” |  |  |
| Neither agree nor disagree | 1 | 4 |
| Agree | 10 | 40 |
| Strongly Agree | 14 | 56 |
| “The materials were easy to understand while providing sufficient detail.” |  |  |
| Neither agree nor disagree | 2 | 8 |
| Agree | 9 | 36 |
| Strongly agree | 14 | 56 |
| “Similar materials could assist conversations about precision medicine application in any disease among patients and general public” |  |  |
| Neither agree nor disagree | 2 | 8 |
| Agree | 13 | 52 |
| Strongly agree | 10 | 40 |

Participant feedback on specific format of IEC materials varied (Table 3). For length, most participants felt the length of both the pamphlet and video was appropriate, though 8% more participants rated the video as “too long” compared to the pamphlet. In terms of visual engagement, 72% agreed or strongly agreed that the pamphlet visuals were engaging, compared to 44% for the video (p = 0.040). For clarity, a majority of participants found the visuals to be simple and easy to understand (Pamphlet: 88%, Video: 80%). Similarly, 72% found the style and presentation of the pamphlet to be well executed, compared to 60% for the video (p = 0.080). When asked about their overall preference, 68% of participants agreed or strongly agreed that they preferred the pamphlet, while 40% expressed a preference for the video (p = 0.363).

**Table 3:**
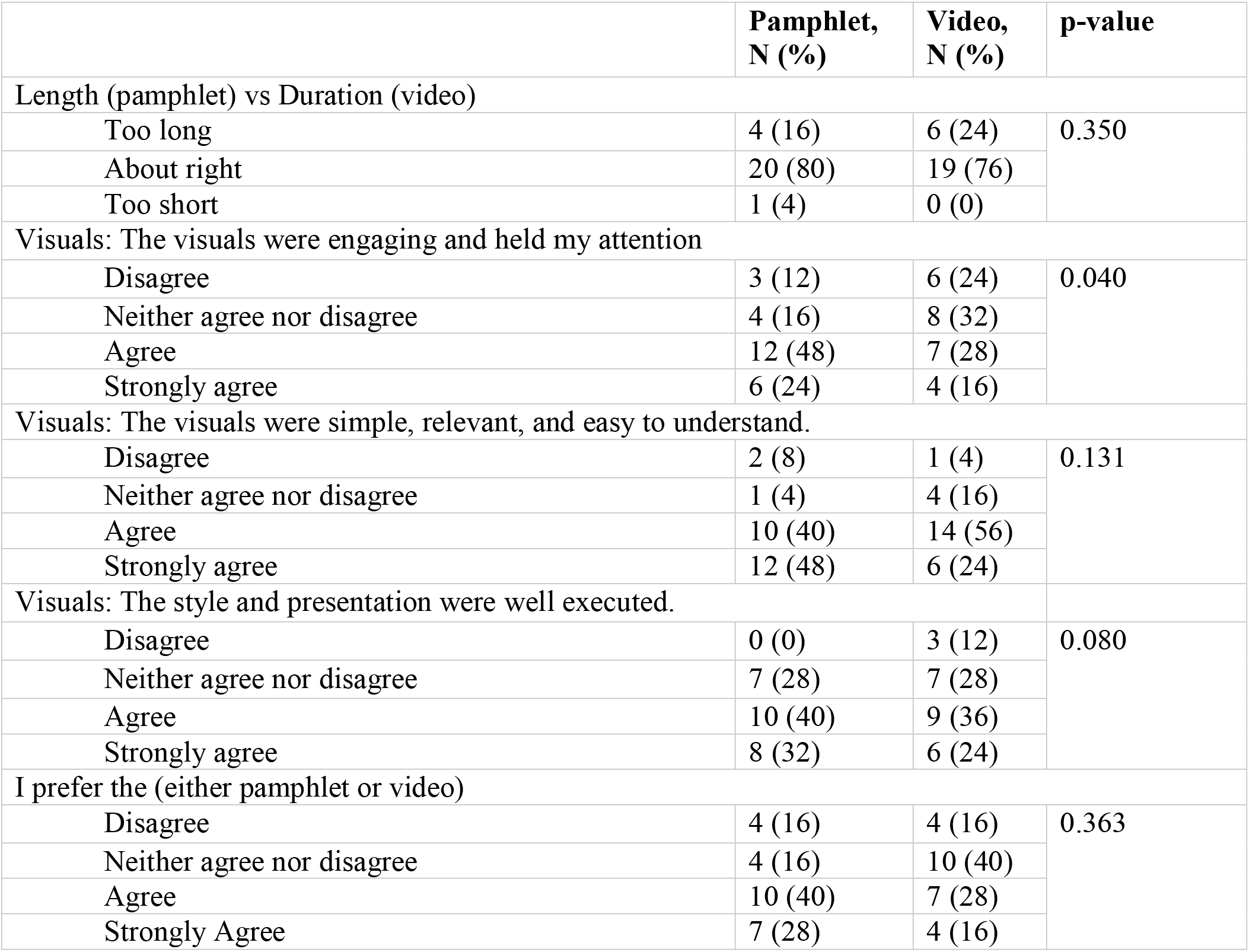
Participant Ratings and Preferences for Pamphlet vs. Video IEC Materials.

Responses to open-ended questions suggested that the materials were complementary and useful when used together. However, participants recommended improving the video content to better illustrate how ACCEPT (prediction tool) functions in clinical scenarios.

## Discussion

The findings of this study offer insights into how complex medical information—particularly prediction tools—can be presented in ways that are accessible and meaningful to diverse audiences. Overall, participants found both the video and pamphlet clear, relevant, and easy to understand. Most found the materials accessible and clear, with 72% indicating they received sufficient information to understand how the tool works and how it could inform treatment decisions. This re-establishes the fact that co-developing materials with patient partners safeguards relevance and clarity, which allows for more patient-centered communication. Furthermore, while both formats were well received, participants preferred the pamphlet due to its concise language and visual clarity. Nevertheless, feedback highlighted the complementary strengths of both formats: the pamphlet supported quick reference, while the video offered narrative context and a patient voice—an approach known to improve emotional engagement and retention.^13^

Quantitative risk prediction tools based on individual genetic, environmental, and lifestyle factors are major enablers of precision medicine.^14^ Despite their promise, their adoption remains limited due to low awareness among patients, providers, and the general public.^15–17^ A content analysis of U.S. print media found that coverage of precision medicine was largely limited to cancer, with minimal attention to its broader applications.^18^ This reinforces the need for improved public education and communication, specially in the field of chronic respiratory diseases such as COPD which is a leading cause of mortality and morbidity worldwide.^19^

One major challenge in communicating prediction tools is low health literacy and numeracy, both essential for interpreting probabilistic risk.^20^ Public-facing materials such as ACCEPT also require a basic understanding of disease management. Information overload and technical jargon can overwhelm patients, impair decision-making, and erode trust in healthcare systems.^21,22^ Understanding this importance, our intervention research design focused on accessible, plain-language IEC resources.

This study has limitations, including a small sample size and participation limited to individuals in the health sciences, which is not reflective of the broader patient population or general public. Nonetheless, it offers early insights into how precision medicine concepts—particularly risk prediction tools—can be communicated in clearer, more accessible ways. The findings will help guide the future development of patient and public centered IEC resources that support the tool’s effective scale-up and integration into routine COPD care and beyond.

## Data Availability

All data produced in the present study are available upon reasonable request to the authors

## Acknowledgement

We gratefully acknowledge the Centre for Lung Health for their support in facilitating our collaboration with the patient-partner through their Community Stakeholder Committee.

## Contributors

JM and SZ are the guarantor of the content of the manuscript, including the data and analysis. Conceptualization (JM and SZ), literature review (JM and SZ), methodology (JM), formal analysis (JM), first drafting of the manuscript (JM and SZ), validation (all authors), review, editing and interpretations (all authors). All authors agreed to be accountable for all aspects of the work.

## Prior Presentations

The article was presented at the ISPOR 2025, May 13-May 16 at Montreal, Canada.

## Notes

### Competing Interest Statement

The authors have declared no competing interest.

### Funding Statement

This study did not receive any funding

### Author Declarations

Ethics committee/IRB of the University of British Columbia gave ethical approval for this work (H23-00445).

