## Supplementary figures and images for "Optimizing Communication Strategies for COPD Management: Effectiveness of Educational Video and Pamphlet Interventions"

### Supplementary Materials

**Supplementary Material:**

**Figure A1: IEC Material (Pamphlet)**

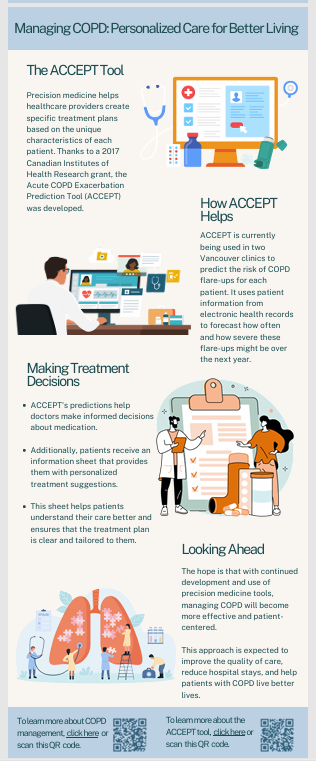

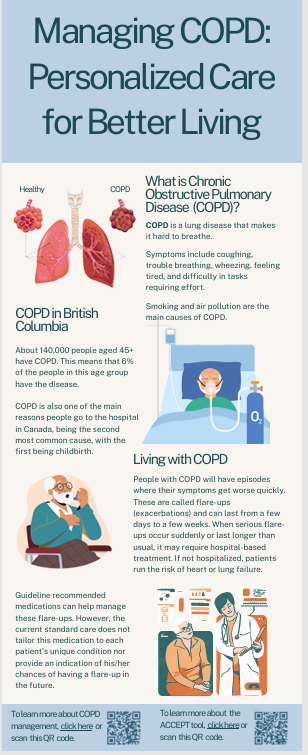
